# A cross ancestry genetic study of psychiatric disorders from India

**DOI:** 10.1101/2024.04.25.24306377

**Authors:** Bharath Holla, Jayant Mahadevan, Suhas Ganesh, Reeteka Sud, Meghana Janardhanan, Srinivas Balachander, Nora Strom, Manuel Mattheisen, Patrick F Sullivan, Hailiang Huang, Peter Zandi, Vivek Benegal, YC Janardhan Reddy, Sanjeev Jain, cVEDA collaborators, ADBS-CBM consortium, iPSYCH OCD consortium, Consortium NORDiC OCD & Related Disorders, Meera Purushottam, Biju Viswanath

## Abstract

Genome-wide association studies across diverse populations may help validate and confirm genetic contributions to risk of disease. We estimated the extent of population stratification as well as the predictive accuracy of polygenic scores (PGS) derived from European samples to a data set from India. We analysed 2685 samples from two data sets, a population neurodevelopmental study (cVEDA) and a hospital-based sample of bipolar affective disorder (BD) and obsessive-compulsive disorder (OCD). Genotyping was conducted using Illumina’s Global Screening Array.

Population structure was examined with principal component analysis (PCA), uniform manifold approximation and projection (UMAP), support vector machine (SVM) ancestry predictions, and admixture analysis. PGS were calculated from the largest available European discovery GWAS summary statistics for BD, OCD, and externalizing traits using two Bayesian methods that incorporate local linkage disequilibrium structures (PGS-CS-auto) and functional genomic annotations (SBayesRC). Our analyses reveal global and continental PCA overlap with other South Asian populations. Admixture analysis revealed a north-south genetic axis within India (*F_ST_* 1.6%). The UMAP partially reconstructed the contours of the Indian subcontinent.

The Bayesian PGS analyses indicates moderate-to-high predictive power for BD. This was despite the cross-ancestry bias of the discovery GWAS dataset, with the currently available data. However, accuracy for OCD and externalizing traits was much lower. The predictive accuracy was perhaps influenced by the sample size of the discovery GWAS and phenotypic heterogeneity across the syndromes and traits studied. Our study results highlight the accuracy and generalizability of newer PGS models across ancestries. Further research, across diverse populations, would help understand causal mechanisms that contribute to psychiatric syndromes and traits.

## INTRODUCTION

Psychiatric genomics has witnessed tremendous growth over the past two decades CITE. The advent of genome-wide association studies (GWAS), along with the formation of international consortia and biobanks, have led to a plethora of genetic associations for psychiatric disorders and brain traits. The success of GWAS has also led to the development of multiple analysis methods to elucidate biological and clinical relevance of these findings (1).

Polygenic score (PGS) is one such method that predicts an individual’s inherited risk for a trait, and is derived as a weighted sum of genomic risk alleles (2) based on GWAS findings. While GWAS requires large sample sizes to detect common variants of small effects, the PGS can be calculated at the level of individuals. Therefore, PGS can be useful for stratifying individuals, for risk of disease, risk based on their genomic variants (3).PGS has gained popularity as a predictive tool for various common disorders. For instance, in coronary artery disease, PGS can identify individuals at increased risk, and the predictive power could be increased even further by combining PGS with clinical risk factors (4–6). In psychiatry, although the discriminative power of PGS is not yet suitable for clinical application, the latest GWAS on schizophrenia has demonstrated promising outcomes, revealing a notably high odds ratio (OR) of 39 (CI 29-53) when comparing the case/control ratio between the highest and lowest deciles of the PGS (7).

However, the under-representation of several global populations represents a major limitation in psychiatric genomics and PGS research. Recent estimates from GWAS of schizophrenia, bipolar disorder (BD) and major depressive disorder suggest that individuals of South Asian ancestry make up less than 1% of these samples (8). The Eurocentric nature of GWAS may limit the optimal transferability of PGS, and may thus restrict the extent to which the genetic discoveries can be extrapolated across diverse populations. As a case in point, the largest meta-analysis from the PGC Schizophrenia Working Group included only 25% participants of non-European ancestry (7). Further, the use of PGS, derived mainly from European-based GWAS, may reinforce or even widen existing health disparities when applied in clinical settings (9). There is now accumulating evidence to suggest that the use of multi-ancestry PGS may perform better than single-ancestry PGS (10). Hence, addressing the lack of ancestral diversity is critical for finessing our findings; as also for an equitable distribution of its potential benefits.

Despite its large share in the global population, the contribution to psychiatry GWAS from India has been sparse. Given the complex population history (11–13), population substructures resulting from geography, linguistics and the historical practice of endogamy (14), there has been a reluctance to apply GWAS and PGS methods in India. However, the advent of large imputation panels and globally representative genotyping arrays, studies have demonstrated feasibility and potential utility of polygenic approaches in India (14). A GWAS on schizophrenia from southern India identified a genome-wide significant locus modifying the expression of *NAPRT1* (15). GWAS in other complex diseases such as diabetes (16) and dyslipidemia (17) have implicated loci, which were previously identified in other continental populations.

This study investigates the predictive accuracy of cross-ancestry PGS for psychiatric phenotypes.. We use state-of-the-art Bayesian methods and also incorporate functional genomic annotations to improve portability from European discovery GWAS. We evaluate the predictive accuracy for BD, OCD, and externalizing symptoms. Moreover, we assess population stratification and admixture levels within these Indian samples, to inform future sampling strategies for Indian ancestry GWAS.

## METHODS

### Samples and phenotypes

Individuals diagnosed to have BD or OCD through the clinical services at NIMHANS were identified, and DNA samples were collected, after informed consent. The cVEDA sample consisted of community samples, identified at random, as part of an ongoing study (ref here)Control subjects were volunteers from the general population, with no lifetime history of psychiatric disease in themselves, or their first degree relatives. All subjects provided written informed consent, and the studies were approved by the ethics committee at NIMHANS.

We used genetic data of consenting individuals from the general population and also patients with OCD or BD. Individuals diagnosed with psychiatric disorders, specifically BD and OCD were identified through clinical services at NIMHANS, Bengaluru, India. All diagnoses were independently confirmed by two experts through detailed clinical interviews and review of medical records, as per the ICD-10 criteria for BD (codes F30 and F31) and OCD (code F42). Most BD patients met the criteria for BD1 syndrome as defined by the DSM5-TR; while those with OCD patients had a moderate to severe illness according to the Clinical Global Impressions Scale.

Individuals without a lifetime psychiatric diagnosis or first-degree family history of psychiatric illness were included as controls along with cVEDA participants. The cVEDA, is a longitudinal study investigating the effects of environmental factors and genomic variations on neurodevelopment and susceptibility to externalizing disorders. The study participants have undergone assessments that covered socio-demographics, temperament, environmental exposures, parenting styles, psychiatric morbidity, and cognitive functioning (18,19). For this study, we focused on the externalizing score from the Strengths and Difficulties Questionnaire (SDQ) CITE, which measures hyperactivity/inattention and conduct problems.

### Genotyping, quality control and imputation

Genotyping was conducted in three batches using the Illumina Global Screening Array, followed by batch-specific quality control using a customized pipeline based on Plink 1.9 and KING version 2.3.2. Samples with genotyping rate < 0.95, F_heterozygosity > 0.2, and genotype-phenotype sex mismatches were removed from the analysis. SNPs were filtered out if these were monomorphic, or had a call rate < 0.95, or deviated from Hardy-Weinberg equilibrium p < 1e^-6^. Pairs of related individuals were identified using the --related function in KING and all first, second, and third degree relatives were excluded. After quality control, the genotypic data of each batch were as follows: Batch1-GSAv2+ had 996 subjects and 619,022 SNPs, Batch2-GSAv3 had 915 subjects and 545,877 SNPs, and Batch3-GSAv3+ included 774 subjects with 633,293 SNPs. Figure S2 illustrates a consistent ∼81% overlap in SNP intersections across the three batches, maintained both pre- and post-QC. A detailed description of genotyping and quality control is provided in the supplementary material.

Following quality control, genotype data was phased using Eagle v2.4 and unobserved genotypes were imputed using Minimac4 with four reference panels: Trans-Omics for Precision Medicine (TOPMed) version r2, Haplotype Reference Consortium (HRC) Version r1.1 2016, 1000 Genomes Phase 3 (Version 5) and the Genome Asia Pilot v1 – (GAsP) on the Michigan Imputation Server. The imputation accuracy was evaluated using info scores (r^2^) across a range of allele frequencies for each reference panel. Following this, “hard call” genotypes from the top-performing panel were filtered for an imputation info score (r^2^) > 0.3 and a minor allele frequency > 0.01. The filtered genotypes were merged for further analysis, resulting in 7,325,802 markers.

### Principal Component Analysis (PCA)

To examine population structure, we conducted PC analysis at three levels: global PCA in conjunction with 1000 Genomes (1000G), continental PCA with Human Genome Diversity Project (HGDP) and Genome Asia Pilot (GAsP), and within-sample PCA. Prior to conducting PCA, the genotype data were pruned for linkage disequilibrium (LD) using an r^2^ of 0.2 and window size of 50 kb and step size of 5 variants. In addition, 24 autosomal regions exhibiting long-range LD and spanning more than 2 Mb were excluded because long-range LD regions (e.g., the extended MHC region) can unduly influence PCA, which can lead to the masking of more subtle genome-wide variation patterns, particularly in admixed samples. This resulted in a final set of 759,439 markers.

For the global and continental PCAs, we merged our samples with publicly available reference datasets. While merging, SNPs with unambiguous allele labels were auto-flipped, while those with ambiguous (A/T, C/G) or inconsistent allele labels were excluded. We also removed any first-, second- and third-degree relatives present in the reference data. The global PCA used 1000G data, including 631 African, 330 Amerindian, 494 East Asian, 500 European, and 454 South Asian samples. Two subsequent Asian sub-continental PCAs (HGDP and GAsP) were conducted to examine how our sample clusters within these sub-continental population structures.

### Ancestry Prediction

We implemented a SVM-based method to identify the most likely ancestral groups for each individual in our sample based on the first 10 global PCs and known ancestry from the 1000 Genomes Project data. The R package ‘e1071’ was used for SVM. An initial parameter grid for the radial kernel SVM was defined in terms of cost and gamma parameters. We used 5-fold cross-validation, repeating the process twice to identify the optimal hyperparameters. Using the best parameters, an SVM model was trained and employed to predict the most probable ancestral group of each individual. The predicted ancestral groups were assigned along with associated probabilities.

### Uniform Manifold Approximation and Projection (UMAP)

The UMAP algorithm was implemented in Python, utilizing the UMAP-learn library, configured with Euclidean distance metric, a neighborhood size of 15, and a minimum distance of 0.1. The first 15 PCs from within-sample PCA were selected for UMAP analysis. The resulting low-dimensional embeddings were plotted with the SVM predicted ancestry labels from the previous step to visualize the within-sample population structure.

### Admixture

Admixture proportions were assessed using ADMIXTURE (v1.3.0). The analysis was performed on genotype data that had been pruned for LD (see above). In alignment with prior literature CITATION, we employed a larger number of markers (272,963) for analysis to achieve effective population resolution, given that the required marker count is inversely proportional to the genetic distance (F_ST_) between the populations under study. The optimal number of ancestral components (K) was determined based on the lowest 10-fold cross-validation error. The algorithm for ADMIXTURE employs maximum likelihood estimation, block relaxation for iterative parameter updates, and a quasi-Newton method for accelerated convergence. A subset of samples with high-confidence self-reported geocodes were plotted on the map of India to examine the geographical distribution of ancestry proportions.

#### Polygenic scores (PGS)

Cross-ancestry PGS were constructed for BD, OCD, and externalizing symptoms using public GWAS summary statistics from European ancestry populations. The discovery cohorts, of individuals of confirmed European ancestry, included 41,917 BD cases and 371,549 controls from the 2021 PGC3 analysis (20); and another 6,848 OCD cases and 18,812 controls from the OCD PGC GWAS (21). For the broad externalizing phenotype, we utilized summary statistics from the genomic structural equation modeling (Genomic SEM) with 1,045,957 participants in the Externalizing Consortium (excluding 23andMe samples as required by this company) (22). For the PGS calculation, we employed three methods: classical clumping and thresholding (C+T) via PRSice-2 (23) and the Bayesian methods PGS-CS-auto (24) and SBayesRC (25). For PRSice-2, the best performing PGS was calculated at a range of P-value thresholds (0.001,0.05,0.1,0.2,0.3,0.4,0.5,1) with default parameters for clumping with a window of 250 kb and r^2^ of 0.1. PGS-CS-auto utilizes a Bayesian regression model with continuous shrinkage priors for posterior SNP effect inference and is robust to different genetic architectures. It models local LD structures using an external reference panel of around 1 million HapMap3 markers. SBayesRC incorporates an LD panel with about 7 million imputed common SNPs along with 96 functional genomic annotations. It employs a hierarchical multi-component mixture prior to adjusting annotation information, thereby influencing the probability and effect size of a SNP being causal. PGS-CS-auto and SBayesRC estimate all parameters jointly within a Bayesian framework, eliminating the need for parameter tuning.

#### PGS Predictive performance

To evaluate the predictive performance of the PGS, we employed a comprehensive set of metrics, with adjustments for covariates including sex, SNP batch, and the first seven principal components of the genotype data. These metrics included: 1) Nagelkerke’s R^2^ (for BD and OCD) and adjusted R^2^ (for externalizing symptoms); 2) Area under the receiver operating characteristic curve (AUC) was calculated for BD and OCD to evaluate the discriminatory power of the PGS; and 3) ORs were estimated to compare the risk between the top 10% of individuals for a given PGS, to the bottom and middle 10% of the sample.

### Multi-PGS Association Analysis

To assess the specificity of associations for BD, OCD and externalizing symptoms, we also attempted an exploratory multi-PGS association analysis with over 3,800 PGSs spanning 18 trait categories, defined according to the Experimental Factor Ontology (EFO) in the PGS Catalog (26). These scores were calculated with ‘pgs-calc’ using default parameters without adjusting for ancestral differences in linkage disequilibrium (LD) or minor allele frequency (MAF). A Bonferroni-corrected significance threshold was set to *α* = 1.3 × 10^−5^ for this analysis, conservatively correcting for total number of PGS tested.

## RESULTS

Following quality control and imputation, the final sample available for subsequent analyses consisted of 2,685 unrelated individuals. These included those with BD (N=463; 47% female), OCD (N=554; 40% female), other psychiatric diagnoses (N=154; 32% female), and neurotypical community controls (N=1514; 48.5% female). The SDQ data on 963 individuals showed a mean Externalizing score of 5.9 (SD=3.6.

As expected, imputation accuracy improved with increasing allele frequency for Indian samples across all four imputation reference panels: HRC, TOPMed, GAsP1, and 1000G (see Supplementary Fig 1). HRC demonstrated the highest accuracy, followed by TOPMed and GAsP1, while 1000G showed the lowest accuracy.

### Population Structure

We employed PCA, UMAP, and ADMIXTURE for assessment of population structure in our study. Global PCA with 1000G data showed that the Indian samples positioned between European and East Asian clusters, with overlap on all the South Asian population labels (Fig 1A). We also observed intra-regional variability, extending along the PC1 axis but condensed along PC2.

**Figure 1:**
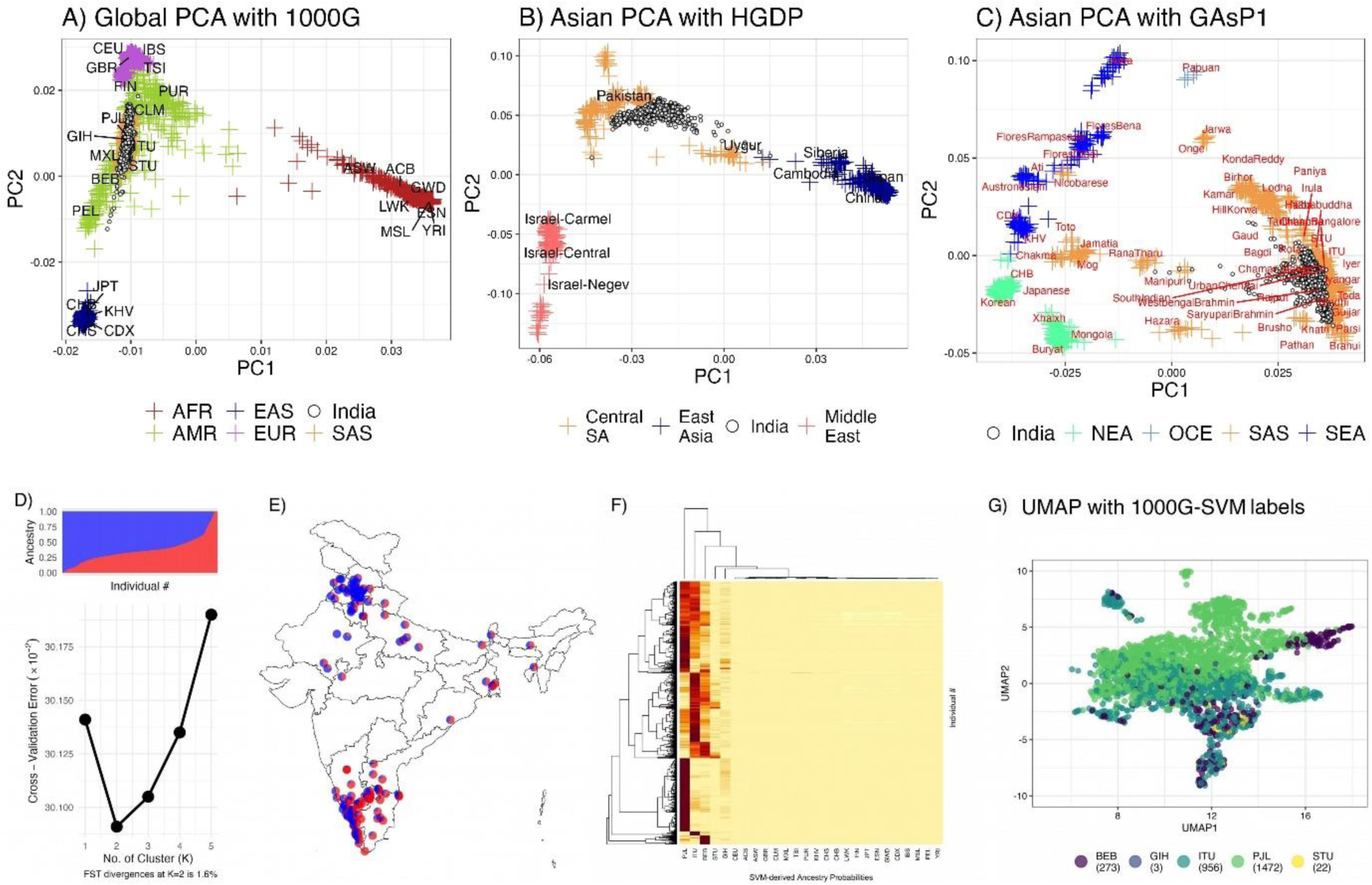
Population structure and genetic diversity in Indian samples. Legend: Global PCA (1A) illustrates the placement of Indian samples between European and East Asian clusters, highlighting notable regional variability. Asian PCA with HGDP samples (1B) reveals distinct clustering of Indian samples, absent in HGDP, situated between Pakistani and Uyghur populations. Asian PCA with Genome Asia samples (1C) demonstrates significant overlap with urban South Asians, indicating urban genetic diversity. ADMIXTURE analysis (1D) reveals a two-cluster genetic structure with a 1.6% F_ST, reflecting modest diversity. Geographic mapping of a subset of samples (1E) presents a north-south ancestral gradient, correlated with latitude. SVM predictions (F) show clustering around key South Asian populations, while UMAP visualization (G) delineates distinct ancestral clusters corresponding to geographical regions, highlighting spatial genetic patterns in India

The Asian PCA plots reveal two contrasting patterns. In the first plot that incorporates HGDP populations, Indian samples do not directly overlap with any HGDP groups. This may well be due to lack of specific Indian ancestry data in HGDP (Fig 1B). Our samples clustered between the Pakistani and Uyghur (China) populations. PCA with Genome Asia Phase 1 data shows that most of our samples form a cluster, substantially overlapping with urban samples from the Genome Asia Phase 1 data (Fig 1C). Considering Genome Asia’s strategic sampling to capture diversity, this overlap suggests that our samples predominantly reflect the genomic spread among urban South Asian populations.

Admixture analysis identified a two-cluster solution for the dataset, supported by an inflection point at K=2 in the cross-validation error curve (Fig 1D). The F_ST_ between these inferred ancestries was 1.6%, surpassing the standard 1% threshold, indicating a modest level of genetic diversity within the sample. Geographic mapping of a subset of samples with available geocodes indicated that ancestral proportions across India are distributed along a north-south gradient (Fig 1E, correlation with latitude r=0.7), thereby suggesting a spatial alignment of genetic ancestry within the country.

The SVM model robustly predicted ancestral groups, assigning probabilities that revealed significant clustering around the PJL, ITU, and BEB populations, with notable representation in STU and GIH, while other groups were less represented (Fig F). Using ancestry predicted by SVM with 1000G global principal components, the UMAP visualization (Fig 1G) approximates the geographical distribution of ancestries in India. Notably, the plot delineates distinct clusters corresponding to Eastern (BEB), Northern (PJL), and Southern Indian (ITU, STU) ancestry groups, which align closely with their respective geographical origins.

### Polygenic Prediction of Psychiatry Phenotypes

We assessed the predictive accuracy of PGS models for BD, OCD, and externalizing traits using PRSice-2, PGS-CS-auto, and SBayesRC. The patterns across the PGS models for BD, OCD, and externalizing traits were consistent (as expected) with imputed data yielding higher predictive accuracy than genotyped data (Figure 2). Notably, Bayesian methods (PGS-CS-auto and SBayesRC) outperformed the classical C+T approach (PRSice-2).

**Figure 2:**
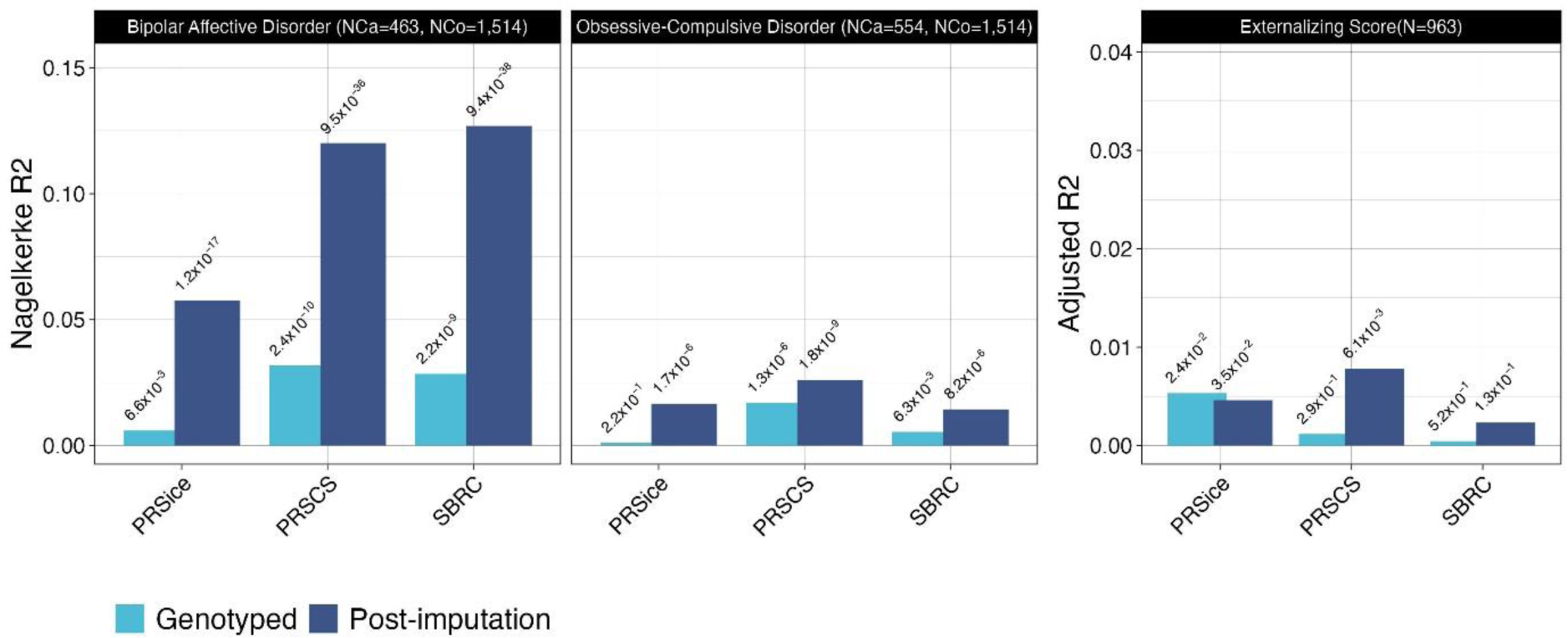
Cross-ancestry polygenic score model comparison for psychiatric phenotypes. Legend: Comparison of European-GWAS derived PGS predictive accuracy for BD, OCD, and externalizing traits in an Indian cohort using PRSice-2, PGS-CS-auto, and SBayesRC. Imputed data shows enhanced prediction, with Bayesian methods outperforming PRSice-2 in cross-ancestry application.

For BD, the application of PGS yielded statistically significant results using both Bayesian methodologies. The p-values were highly significant at 9.49×10^−36^ for PGS-CS-auto and 9.38×10^−38^ for SBayesRC, accompanied by Nagelkerke R^2^ values of 0.120 and 0.127, respectively. Furthermore, when assuming a 1% population prevalence, these PGS models explain approximately 6.5% (using PGS-CS-auto) and 6.9% (using SBayesRC) of the phenotypic variance on the liability scale. The discriminatory ability of these models is substantiated by an AUC of around 71% for both methods. In terms of clinical risk stratification, the PRS-CS-auto method yields an ORs of 3.5 (95% CI 2.2–5.6) and 12.8 (95% CI 6.4–25.3) for the individuals in the top decile of being affected with the disorder, when compared to individuals in the middle and lowest deciles, respectively (Figure 3). In contrast, SBayesRC yields higher corresponding ORs of 3.8 (95% CI 2.4–5.9) and 20.5 (95% CI 9.4–44.4).

**Figure 3:**
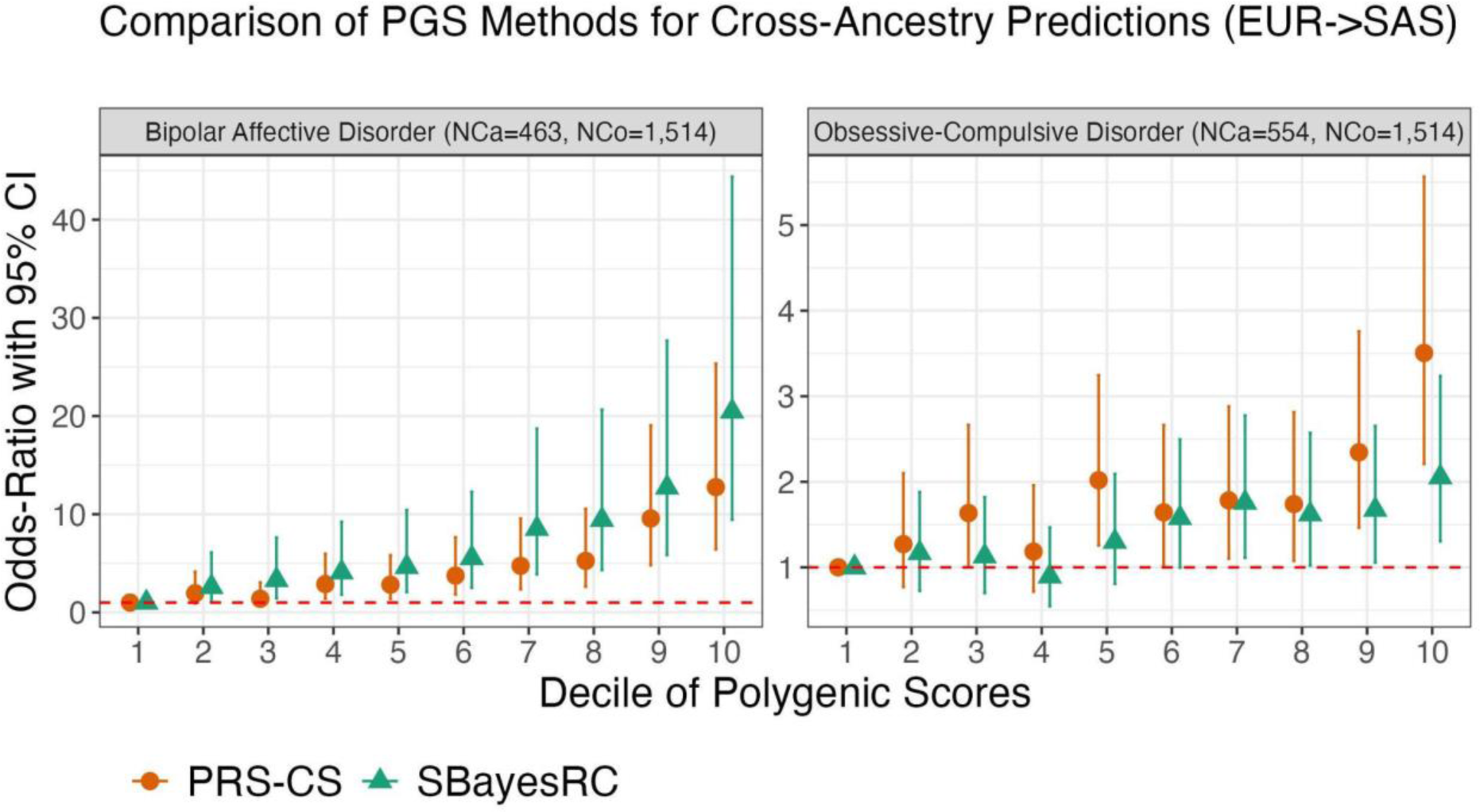
Decile-based risk stratification in BD and OCD. Legend: Both PRS-CS-auto and SBayesRC methods stratify risk across deciles for BD (3A) and OCD (3B), with BD showing greater discriminative power.

For OCD, the application of PGS also showed significant findings using both PRS-CS-auto and SBayesRC methods. The p-values were significant at 1.84×10^-9^ for PRS-CS-auto and 8.23×10^-6^ for SBayesRC, with corresponding Nagelkerke R^2^ values of 0.026 and 0.014, respectively. When assuming a 1% population prevalence, these models account for approximately 1.3% (PRS-CS-auto) and 0.7% (SBayesRC) of the phenotypic variance on the liability scale. The models’ discriminatory ability was lower compared to BD, with an AUC of approximately 59% for both methods. In terms of clinical risk stratification for OCD, PRS-CS-auto yields odds ratios of 2.2 (95% CI 1.4–3.4) and 3.5 (95% CI 2.2–5.6) individuals in the top and lowest deciles, respectively, as compared to those in the middle decile. SBayesRC demonstrates lower odds ratios of 1.3 (95% CI 0.9–2.1) and 2.1 (95% CI 1.3–3.2).

For externalizing symptoms, the PRS-CS-auto model accounted for approximately 0.78% of the variance in the trait, with a coefficient estimate of 0.094 (95% CI: 0.027-0.160, p-value = 0.006). In contrast, the SBayesRC method explained a smaller variance of 0.23% and a coefficient estimate of 0.057 (95% CI: −0.017-0.130, p-value = 0.133).

## Discussion

In this study, we evaluated the predictive accuracy of PGS derived from European ancestry populations as applied to an Indian sample. We also evaluated the genetic structure of several samples ascertained in India with the intent of assessing its genetic diversity, both globally and regionally.

We identified a robust cross-ancestry overlap in BD PGS. Our sample included only those with well-characterized BD1 and high heritability. This hints at considerable overlap in underlying biological mechanisms across ancestries. Interestingly, OCD PGS also showed a significant cross-ancestry prediction, albeit with smaller effect size compared to BD likely due to the far smaller number of cases in the European OCD GWAS. Overall, our analyses demonstrate that PGS models, developed from European cohorts and applied to Indian samples using Bayesian methodologies that adjust for local LD patterns and incorporate functional genomic annotations, maintain statistical significance for psychiatric diagnoses. These findings suggest a shared polygenic liability across populations, indicating that the genetic foundations of psychiatric disorders remain consistent, and are detectable, across diverse ethnic backgrounds (27).

The PGS for externalizing symptoms exhibited lesser overlap than that found for BD and OCD. This difference could be attributed to the methods of phenotypic ascertainment. Individuals with BD and OCD were identified through the clinical services, evaluated using structured face-to-face interviews, and diagnosed using well established and reliable diagnostic guidelines. Those with externalizing traits are typically gauged through standardized questionnaires in varied settings, resulting ina lower diagnostic accuracy. Moreover, the inherent genetic attributes of each disorder may impact the effectiveness of PGS. For example, BD may present more consistent genetic signals, on account of its greater heritability, when compared to externalizing traits, which are likely influenced by a wider array of genetic and environmental factors.

The genetic diversity observed in our sample is broadly comparable to that seen in European populations, which exhibit variation along both East-West and North-South axes (28,29). The investigation of genetic risks for common disorders, prevalent across all populations, underscores the necessity of including large, continental samples. Despite these encouraging findings, it may still be useful to assess the impact of specific patterns of diversity, within sub-populations, on these risks. Such approaches promise to enhance the generalizability of genetic models in research, targeting the genetic underpinnings of common medical conditions The lack of global diversity in genetic studies impacts the generalizability of findings for underrepresented populations, and constrains the understanding of genetic bases of traits across varied groups. This is relevant in psychiatric disorder research, which has predominantly included individuals of European ancestry to identify associated genetic loci. The epidemiology, and semiology, of psychiatric syndromes such as BD1 and OCD are quite similar all over the world. This suggests that there is likely a similar genetic liability, with a shared set of genomic loci influencing these phenotypes across populations (30). Nonetheless, ancestry-related variations in LD patterns, with the true causal variant, as well as diverse environmental interactions, may affect the size of the genetic effect.

Our results demonstrate the promising portability of PGS based on European GWAS for South Asian samples. The genetic variation between populations, manifested in LD structure and allele frequency differences, provides opportunities to discover new loci linked to psychiatric disorders. Such differences in LD structure are instrumental in fine-mapping associated regions, helping to close in on causal variants responsible for the observed associations. Our findings underline the need for further refinement of these models through more diverse and inclusive multi-ancestral research efforts. Initiatives like the Ancestral Population Network studies (https://www.nimh.nih.gov/about/organization/dnbbs/genomics-research-branch/ancestral-populations-network-apn), including the Asian Bipolar Genetics Network (A-BIG-NET) (31), are pivotal in promoting genetic diversity within psychiatric genetics research. Such efforts are expected to deepen our understanding of the genetics underlying psychiatric conditions, highlighting the shared and unique aspects of psychiatric phenotypes across different populations.

## Appendices

1. Supplementary Material (see below)
  1. Figures
  2. Tables
2. Data Availability Statement

## Collaborators

### cVEDA consortium

Jon Heron 1, Matthew Hickman 1, Debashish Basu 2, Subodh Bhagyalakshmi Nanjayya 2, Rajkumar Lenin Singh 3, Roshan Lourembam 4, Kalayanaraman Kumaran 5, Murali Krishna 6, Rebecca Kuriyan 7, Sunita Simon Kurpad 8, Kamakshi Kartik 9, Kartik Kalyanram 9, Sylvane Desrivieres 10, Gareth Barker 11, Dimitri Papadopoulos Orfanos 12, Mireille Toledano 13, Rose Dawn Bharath 14, Pratima Murthy 15, Eesha Sharma 16, Nilakshi Vaidya 17, Amit Chakrabarti 18, Gunter Schumann 17,

1. – Population Health Sciences, Bristol Medical School, University of Bristol, United Kingdom
2. – Department of Psychiatry, Post Graduate Institute of Medical Education and Research, Chandigarh, India
3. – Department of Psychiatry, Regional Institute of Medical Sciences, Imphal, India
4. – Department of Psychology, Regional Institute of Medical Sciences, Imphal, India
5. – MRC Lifecourse Epidemiology Unit, University of Southampton, United Kingdom & Epidemiology Research Unit, CSI Holdsworth Memorial Hospital, Mysuru, India
6. – Foundation for Research and Advocacy in Mental Health, Mysuru, India
7. – Division of Nutrition, St John’s Research Institute, Bengaluru, India
8. – Department of Psychiatry & Department of Medical Ethics, St. John’s Medical College & Hospital, Bengaluru, India
9. – Rishi Valley Rural Health Centre, Madanapalle, Chittoor, India
10. – Centre for Population Neuroscience and Precision Medicine, Institute of Psychology, Psychiatry & Neuroscience, MRC SGDP Centre, King’s College London, United Kingdom 11 – Department of Neuroimaging, Institute of Psychology, Psychiatry & Neuroscience, King’s College London, United Kingdom
11. – NeuroSpin, CEA, Universit’e Paris-Saclay, Paris, France
12. – MRC Centre for Environment and Health, School of Public Health, Imperial College, London, United Kingdom
13. – Department of Neuroimaging and Interventional Radiology, National Institute of Mental Health and Neurosciences, Bengaluru, India
14. – Department of Psychiatry, National Institute of Mental Health and Neurosciences, Bangalore, India
15. – Department of Child and Adolescent Psychiatry, National Institute of Mental Health and Neurosciences, Bangalore, India
16. – Centre for Population Neuroscience and Precision Medicine, Charite Mental Health, Dept. of Psychiatry and Psychotherapy, Charite Universitaetsmedizin Berlin, Germany;
17. Centre for Population Neuroscience and Precision Medicine, Institute for Science and Technology of Brain-Inspired Intelligence, Fudan University, Shanghai, China
18. – Indian Council of Medical Research–Centre for Ageing and Mental Health, Kolkata, India

### ADBS-CBM consortium

Biju Viswanath 1, Ganesan Venkatasubramanian 1, John P John 1, Meera Purushottam 1, Reeteka Sud 1, Bharath Holla 1, Jayant Mahadevan 1, Srinivas Balachander 1, Sreeraj VS 1, G Vijay Kumar 1, Suhas Ganesh 1, Suhas Satish 1, Preethi V. Reddy 1, Vijaykumar S Harbishettar 1, Lekhansh Shukla 1, Pradip Paul 1, Bhagyalakshmi M. S 1, Palanimuthu T Sivakumar 1, Arun Kandasamy 1, Muralidharan Kesavan 1, Urvakhsh Meherwan Mehta 1, Ashitha S. N. M 1, Bhupesh Mehta 2, Thennarasu Kandavel 3, B Binukumar 3, Jitender Saini 4, A Shyamsundar 1, Gautam Arunachal Udupi 5, Himani Kashyap 6, Anish V Cherian 7, K S Meena 8, Latha K 8, Jagadisha Thirthalli 1, Prabha S Chandra 1, Pratima Murthy 1, Upinder S Bhalla 9, Vivek Benegal 1, Padinjat Raghu 9, Janardhan Y C Reddy 1.

1. – Department of Psychiatry, National Institute of Mental Health and Neurosciences, Bangalore, India
2. – Department of Biophysics, National Institute of Mental Health and Neurosciences, Bangalore, India
3. – Department of Biostatistics, National Institute of Mental Health and Neurosciences, Bangalore, India
4. – Department of Neuroimaging and Interventional Radiology, National Institute of Mental Health and Neurosciences, Bangalore, India
5. – Department of Human Genetics, National Institute of Mental Health and Neurosciences, Bangalore, India
6. – Department of Clinical Psychology, National Institute of Mental Health and Neurosciences, Bangalore, India
7. – Department of Psychiatric Social Work, National Institute of Mental Health and Neurosciences, Bangalore, India
8. – Department of Mental Health Education, National Institute of Mental Health and Neurosciences, Bangalore, India
9. – National Centre for Biological Sciences (NCBS)

### iPSYCH OCD Consortium

Thomas Damm Als 1,2,3, Anders D. Børglum 1,2, Nora Strom 4, Manuel Mattheisen 4,5,6, Jonas Bybjerg-Grauholm 2,7, Jakob Grove 1,2,3,8, David M. Hougaard 2,7, Ole Mors 9, Preben B. Mortensen 10,11, Judith Becker Nissen 12,13, Merete Nordentoft 14,15, Thomas Werge 15,16,17

1. – Department of Biomedicine, Aarhus University, Aarhus, Denmark.
2. – The Lundbeck Foundation Initiative for Integrative Psychiatric Research, iPSYCH, Copenhagen, Denmark.
3. – Center for Genomics and Personalized Medicine, Aarhus, Denmark.
4. – Institute of Psychiatric Phenomics and Genomics (IPPG), University Hospital, LMU Munich, Munich, Germany
5. – Dalhousie University, Department of Community Health and Epidemiology & Faculty of Computer Science, Halifax, Nova Scotia, Canada
6. – University Hospital of Psychiatry and Psychotherapy, University of Bern
7. – Department for Congenital Disorders, Statens Serum Institut, Copenhagen, Denmark
8. – Bioinformatics Research Centre, Aarhus University, Aarhus, Denmark
9. – Psychosis Research Unit, Aarhus University Hospital - Psychiatry, 8200 Aarhus N, Denmark
10. – National Centre for Register-based Research, Aarhus University, Aarhus, Denmark
11. – Centre for Integrated Register-based Research, Aarhus University, Aarhus, Denmark
12. – Departments of Child and Adolescent Psychiatry, Aarhus University Hospital, Psychiatry, Denmark, Aarhus University Hospital, Psychiatry, Aarhus, Denmark
13. – Institute of Clinical Medicine, Health, Aarhus University, Health, Aarhus University, Aarhus, Danmark
14. – CORE - Copenhagen Research Center for Mental Health, Copenhagen University Hospital, Copenhagen, Denmark
15. – Department of Clinical Medicine, University of Copenhagen, Copenhagen, Denmark
16. – Department of Mental Health Services, Copenhagen University Hospital, Copenhagen, Denmark
17. – GLOBE Institute, University of Copenhagen, Copenhagen, Denmark

### NORDiC OCD & Related Disorders Consortium (NORDiC)

Julia Boberg 1, Long Long Chen 1, James J. Crowley 1,2, Elles de Schipper 1, Diana R. Djurfeldt 1, Jan Haavik 3,4, Kristen Hagen 3,5,6, Matthew W. Halvorsen 1,2, Bjarne Hansen 3,7, Kira D. Höffler 3, Anna K. Kähler 8, Elinor K. Karlsson 9,10,11, Gerd Kvale 3,12, Paul Lichtenstein 8, Kerstin Lindblad-Toh 9,13, Nora Strom 14, Manuel Mattheisen 14,15,16, David Mataix-Cols 1,17, Kathleen Morrill 10,18, Hyun Ji Noh 9, Christian Rück 1, Thorstein Olsen Eide 5,7, Tetyana Zayats 21,22

1. – Centre for Psychiatry Research, Department of Clinical Neuroscience, Karolinska Institutet & Stockholm Health Care Services, Region Stockholm, Sweden
2. – Department of Genetics, University of North Carolina at Chapel Hill, NC, USA
3. – Bergen Center for Brain Plasticity, Division of Psychiatry, Haukeland University Hospital, Bergen, Norway
4. – Department of Biomedicine, University of Bergen, Bergen, Norway
5. – Department of Psychiatry, Molde Hospital, Molde, Norway
6. – Department of Mental Health, Norwegian University of Science and Technology, Trondheim, Norway
7. – Center for Crisis Psychology, Faculty of Psychology, University of Bergen, Norway
8. – Department of Medical Epidemiology and Biostatistics, Karolinska Institutet, Stockholm, Sweden
9. – Broad Institute of MIT and Harvard, Cambridge, MA 02139, USA
10. – Program in Bioinformatics and Integrative Biology, UMass Chan Medical School, Worcester, MA 01605, USA
11. – Program in Molecular Medicine, UMass Chan Medical School, Worcester, MA 01605, USA
12. – Department of Clinical Psychology, University of Bergen, Norway
13. – Science for Life Laboratory, Department of Medical Biochemistry and Microbiology, Uppsala University, 751 32 Uppsala, Sweden
14. – Institute of Psychiatric Phenomics and Genomics (IPPG), University Hospital, LMU Munich, Munich, Germany
15. – Dalhousie University, Department of Community Health and Epidemiology & Faculty of Computer Science, Halifax, Nova Scotia, Canada
16. – University Hospital of Psychiatry and Psychotherapy, University of Bern
17. – Department of Clinical Sciences, Lund University, Lund, Sweden
18. – Morningside Graduate School of Biomedical Sciences UMass Chan Medical School, Worcester, MA, USA.
19. – Department of Psychology, Humboldt-Universität zu Berlin, Berlin, Germany
20. – Department of Biomedicine, Aarhus University, Aarhus, Denmark
21. – K.G. Jebsen Center for Research on Neuropsychiatric Disorders, Department of Biomedicine, University of Bergen, Bergen, Norway.
22. – Center for Medical Genetics and Molecular Medicine, Haukeland University Hospital, Bergen, Norway.

## Data Availability

All data produced in the present study are available upon reasonable request to the authors

---

**SFigure 1:**
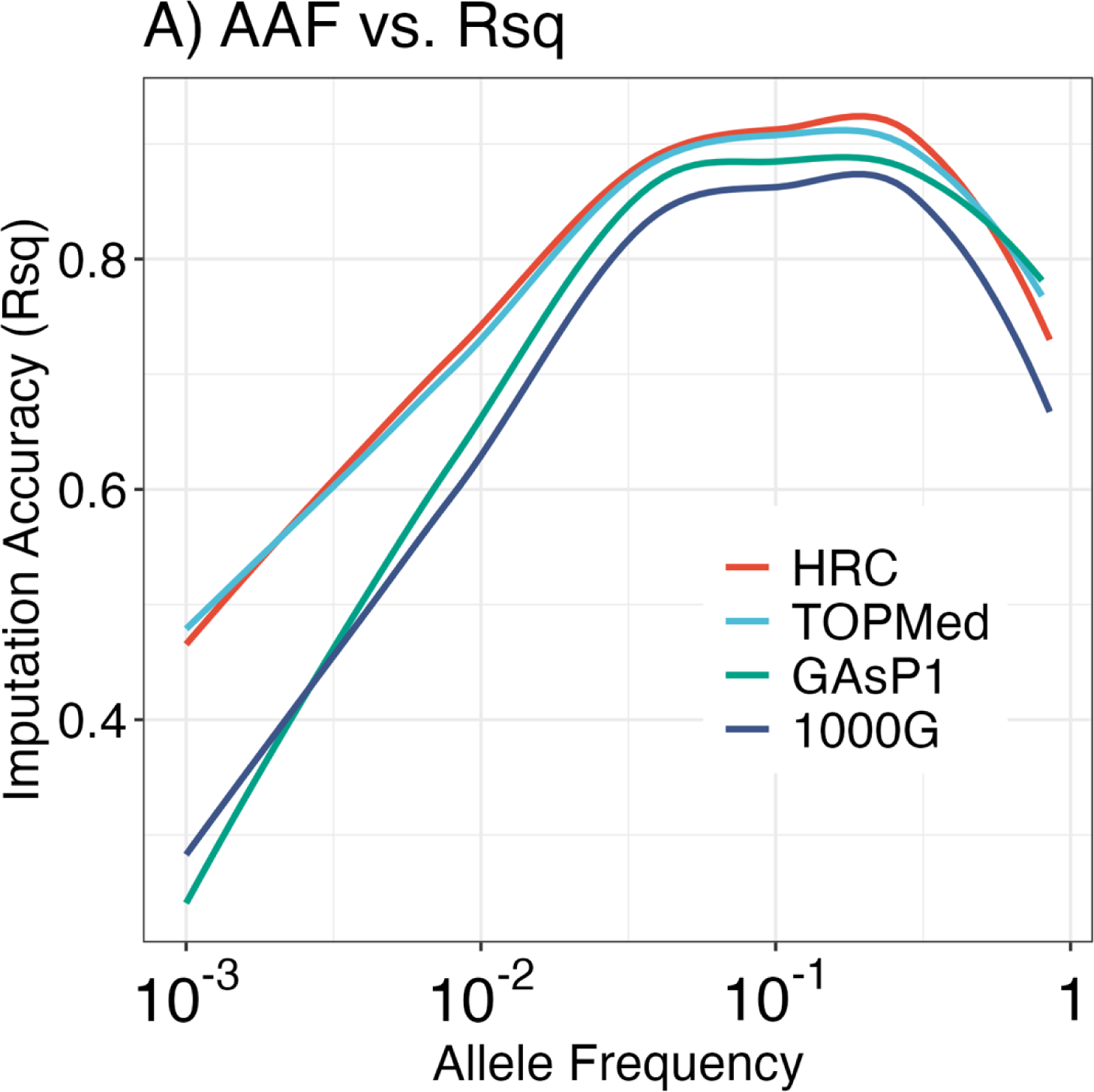
Comparative Imputation Accuracy Across Allele Frequencies on four imputation reference panels: HRC, TOPMed, GAsP1, and 1000G. Imputation with HRC reference panel showed superior accuracy compared to other panels across allele frequency spectrum.

**SFigure 2:**
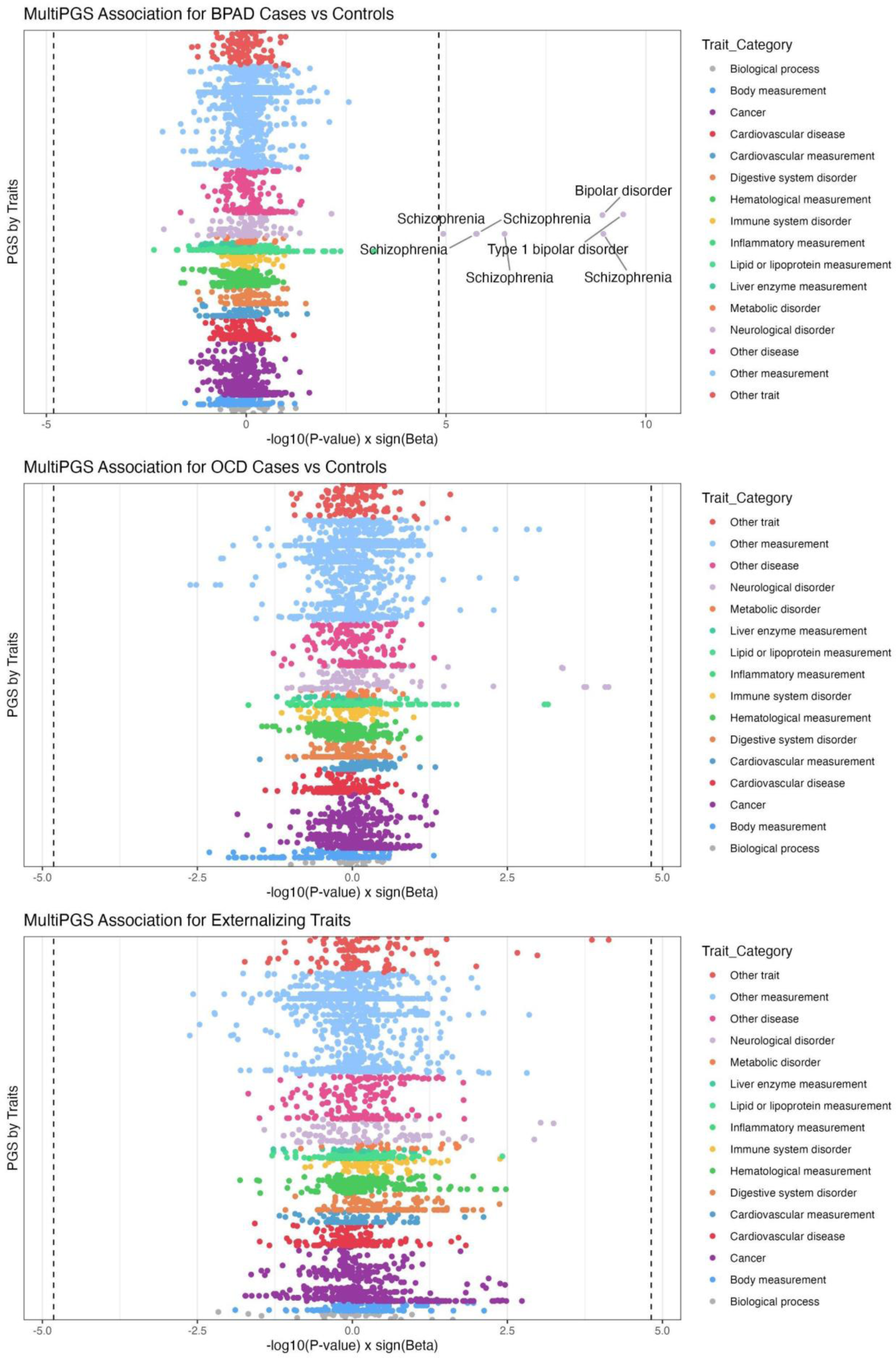
Exploratory Multi-Polygenic Score (MultiPGS) Associations. The plot shows −log10(P-value)×sign(β) for each trait within 18 categories in relation to Bipolar Disorder, Obsessive-Compulsive Disorder, and Externalizing traits. The traits are organized according to the Experimental Factor Ontology (EFO) and referenced in the PGS Catalog. The dotted line signifies the significance threshold adjusted for multiple testing at −log10(0.05/number of tests).

**SFigure 3.**
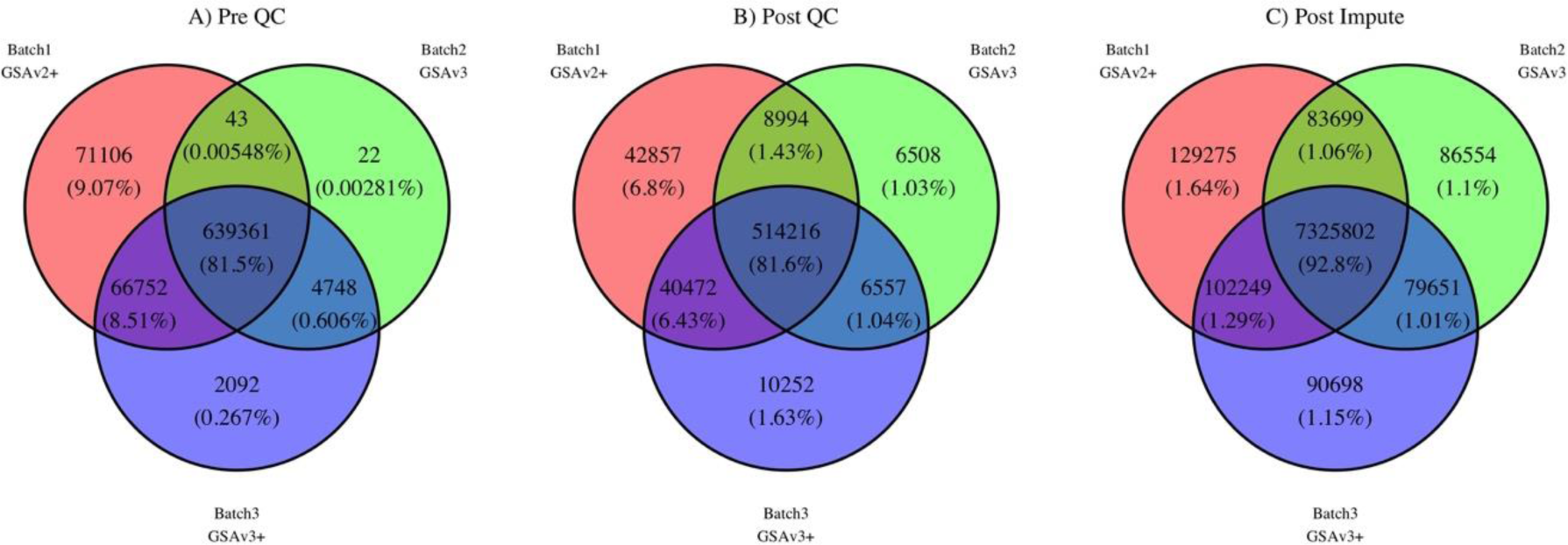
Quality Control of SNP Genotyping Across Different Batches: The Venn diagrams depict SNP intersections across three batches using the Global Screening Array, before (A) and after (B) quality control (QC), and following imputation with the HRC reference panel (C). They reveal a significant overlap of approximately 80-90% in all batches, consistent both pre and post QC and imputation. The ‘+’ symbol denotes batches enhanced with additional custom markers from the Infinium PsychArray-24’s focused content panel

**STable2**: MultiPGS results

